# Does COVID-19 Increase the Risk of Subsequent Kidney Diseases More Than Influenza? A Retrospective Cohort Study Using Real-World Data In the United States

**DOI:** 10.1101/2024.06.27.24309556

**Authors:** Yue Zhang, Nasrollah Ghahramani, Vernon M. Chinchilli, Djibril M. Ba

## Abstract

**Background:** Although case reports and observational studies suggest Coronavirus disease 2019 (COVID-19) increases the risk of kidney diseases, definitive real-world evidence, especially in comparison with influenza, is lacking. Our study aims to assess the association between COVID-19 infections and subsequent kidney diseases, using influenza as a positive control and incorporating a negative control to establish clearer associations.

**Methods:** A large retrospective cohort study with strata matching was conducted using the MarketScan database with records from Jan. 2020 to Dec. 2021. We used the international classification of 10th revision (ICD-10) codes to identify individuals and build three cohorts, (1) COVID-19 group, with index dates as the diagnosis dates of COVID-19; (2) Influenza but no COVID-19 (positive control) group, with index dates as the diagnosis dates of Influenza; and (3) no COVID-19 / Influenza (negative control) group, with randomly assigned index dates between Jan. 2020 to Dec. 2021. The main outcomes were acute kidney injury (AKI), chronic kidney disease (CKD), and end-stage renal disease (ESRD). To evaluate the association between COVID-19 and the new onset of kidney diseases relative to both control groups, we employed multivariable stratified Cox proportional hazards regression analysis.

**Results:** The study included 939,241 individuals with COVID-19, 1,878,482 individuals in the negative control group, and 199,071 individuals with influenza. After adjusting for demographics, comorbidities, and medication histories, COVID-19 was significantly associated with increased risks of AKI (adjusted hazards ratio, aHR: 2.74, 2.61-2.87), CKD (aHR: 1.38, 1.32-1.45), and ESRD (aHR, 3.22; 95% CI, 2.67-3.88), while influenza was associated with a modestly increased risk of AKI (aHR: 1.24, 1.11-1.38) and had no impact on CKD (aHR: 1.03, 0.92-1.14), and ESRD (aHR, 0.84; 95% CI, 0.55-1.29). Time-specific analyses indicated that while the HR for AKI declined from 0-180 days to 0-540 days, the HR for CKD and ESRD remained stable, with COVID-19’s risk surpassing influenza’s risk throughout follow-up. Exploratory analysis also found significant impacts of COVID-19 on glomerular diseases (aHR 1.28, 95% CI 1.09-1.50).

**Conclusion:** In this large real-world study, COVID-19 infections were associated with a 2.3-fold risk of developing AKI, a 1.4-fold risk of CKD, and a 4.7-fold risk of ESRD compared to influenza. Greater attention needs to be paid to kidney diseases in individuals after contracting COVID-19 to prevent future adverse health outcomes.

## Introduction

Coronavirus disease 2019 (COVID-19) has been reported to be associated with multiple short-term and long-term conditions following exposure^1–6^. Kidney diseases, including short-term conditions like acute kidney injury (AKI) and long-term conditions like chronic kidney disease (CKD) and glomerular diseases, have attracted significant research attention, particularly regarding the long-term and short-term effects of COVID-19^7–9^. In addition to studying the effects of COVID-19 compared to non-infected individuals, there is increasing interest in comparing the impact of COVID-19 to influenza, one of the most common viral respiratory illnesses in the US^10^. Studies showed that COVID-19 is associated with higher risks of mortality and several comorbidities, including cardiovascular diseases, mental health disorders, and neurologic diseases, compared to influenza^11–13^. Therefore, a better understanding of the effects of COVID-19, in comparison to influenza, on short-term and long-term kidney diseases is needed to provide more comprehensive information to the public.

Previous studies assessing the associations between COVID-19 and kidney diseases have primarily focused on specific populations, such as U.S. veterans or hospitalized patients, and often lacked comparisons with influenza^7,14–16^. To the best of our knowledge, there is currently no study that has investigated the association among COVID-19, positive controls (influenza only), and negative controls (neither COVID-19 nor influenza infections) and the risk of kidney diseases using a large real-world database. In our study, we included over 3 million individuals from national real-world datasets (MarketScan) in the United States. Additionally, we compared the effects of COVID-19 to both positive control group and negative control group regarding the risks of AKI, CKD, end-stage renal disease (ESRD), and glomerular diseases. We hypothesized that individuals with COVID-19 infections have higher risks of kidney diseases compared to both positive and negative control groups, in both the short term and long term.

## Methods

### Data Source

This retrospective cohort study utilized data from Merative™ MarketScan^®^ Commercial Database (hereafter referred to as MarketScan), ranging from January 2019 to December 2021. MarketScan is one of the largest and most comprehensive longitudinal claims databases available for healthcare research, encompassing data on over 275 million unique de-identified patients^17^. This database includes health insurance claims across various types of care (e.g., inpatient, outpatient, pharmacy, and other claims), as well as enrollment data from large employers and health plans across all 50 U.S. states and the District of Columbia. The longitudinal tracking of detailed patient-level healthcare claims information provides comprehensive data, including demographic characteristics such as age, sex, diagnoses, procedures, and medications^18^.

The included individuals had continuous enrollment for one year prior to, and 90 days following, the index dates. Data from 2020 and 2021 were primarily used for analytical purposes, while one year before the index dates was utilized to define baseline covariates.

### Cohort Derivation and Assessment of Exposure

Individuals with COVID-19, aged between 18 to 64 years, were identified from 1 January 2020 to 31 December 2021, using the International Classification of Diseases, 10th Revision (ICD-10) (*Supplementary Table 1*). The date of the first COVID-19 diagnosis was defined as the individual’s index date. Two control groups were also established: (1) Positive control group: This group consisted of individuals with influenza, identified using ICD-10 code (*Supplementary Table 1*), but no COVID-19 diagnosis histories. The recorded first diagnosis dates served as the index dates; (2) Negative control group: This group included individuals without any history of COVID-19 or influenza diagnoses, and pseudo random index dates were assigned between 1 January 2020 and 31 December 2021.

After excluding individuals with a history of kidney diseases at the study baseline, the COVID-19 group and the negative control group were matched at a maximum ratio of 1:2 across 1,536 strata. These strata were defined by year of birth (ranging from 1956 to 2003), gender (male or female), region (northeast, north-central, south, and west), and index date periods. Matching by index date periods aimed to control for the effects of temporal variations in COVID-19^19^. The periods were defined based on the dominant SARS-CoV-2 variants as follows: (1) January to June 2020: only Alpha (B1.1.7); (2) July to December 2020: Alpha (B1.1.7), Beta (B.1.351) and Gamma (P.1); (3) January to June 2021: Beta (B.1.351) and Delta (B.1.617.2); (4) July to December 2021: Delta (B.1.617.2)^20^.

Strata that did not achieve the exact 1:2 ratio between the COVID-19 group and the negative control group were excluded, resulting in 1,469 strata being included in the final analysis. Individuals with influenza were directly assigned to these existing 1,469 strata (*Figure 1*).

**Figure 1:**
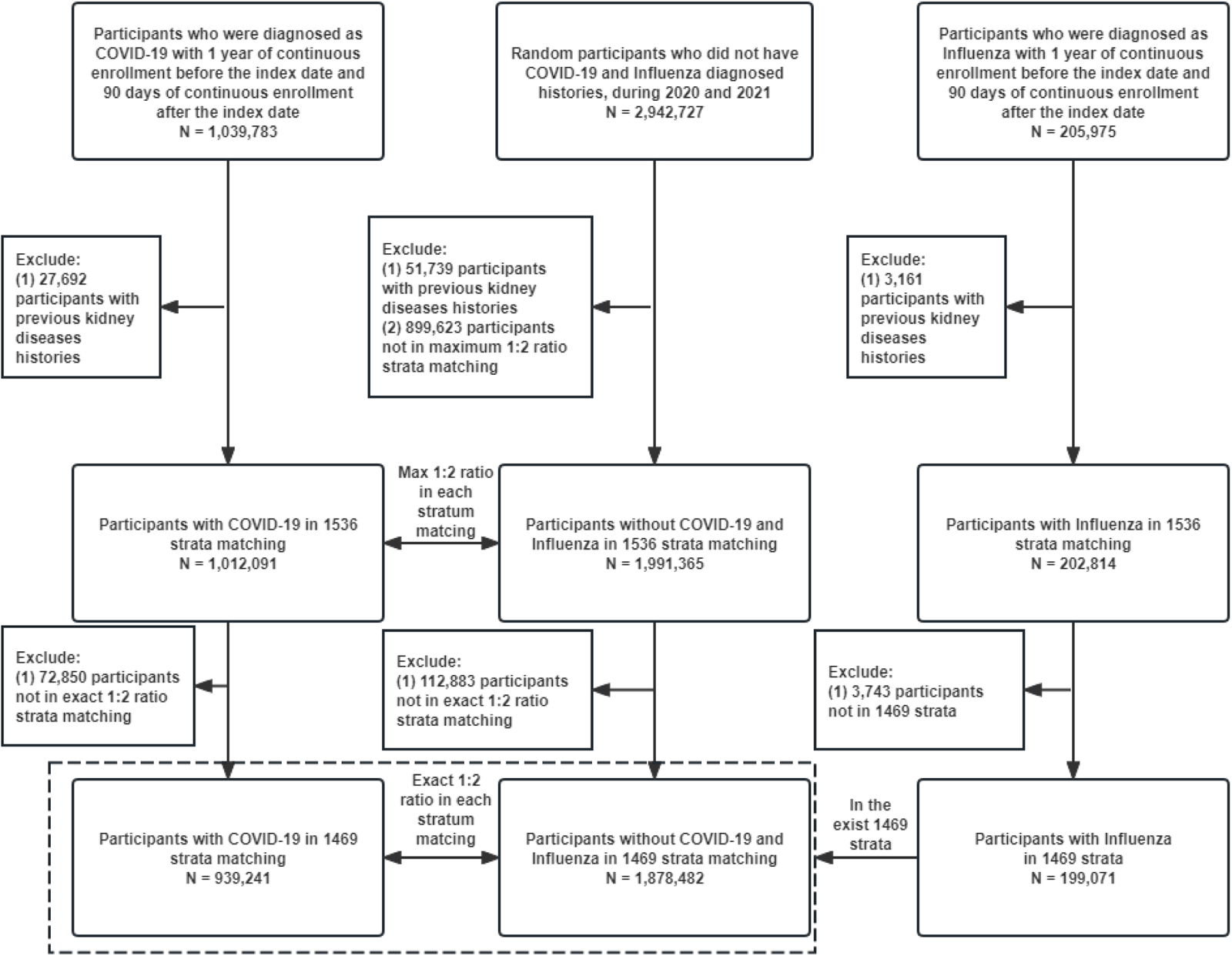
Participant Selection Flow Diagram for COVID-19, Positive Control, and Negative Control Groups from the MarketScan Dataset

### Assessment of Outcomes

The new onset of kidney diseases was assessed during the follow-up period, defined as the time between kidney disease onset and the index date. The analysis was censored at the occurrence of death, kidney disease, or the end of the study (31 December 2021), whichever occurred first. Kidney diseases were identified using ICD-10 codes, with all kidney diseases and subgroups, including AKI, CKD, and ESRD (including dialysis and kidney transplant). Glomerular disease was also identified for the exploratory analysis (*Supplementary Table 1*).

### Assessment of Potential Covariates

Demographic data on age, sex, and US regional location were extracted directly from MarketScan datasets. Individuals’ comorbidities were assessed by the Elixhauser Comorbidity Index within one year prior to the index dates, utilizing ICD-10 codes^21,22^. The six most prevalent conditions, which are potentially related to kidney disease, COVID-19, or influenza, include cardiovascular disease (CVD, including congestive heart failure, cardiac arrhythmia, valvular disease, pulmonary circulation disorders, peripheral vascular disorders, and hypertension)^23–25^, diabetes^26,27^, obesity^28,29^, chronic pulmonary disease^30,31^, liver diseases^32,33^, and depression^34,35^ (*Supplementary Table 1*). In addition, we examined the usage history of seven specific medications – angiotensin-converting enzyme inhibitors or angiotensin II receptor blockers (ACEI/ARBs), statins, diuretics, calcium channel blockers (CCB), nonsteroidal anti-inflammatory drugs (NSAIDs), proton pump inhibitors (PPIs), and beta-blockers – within the year preceding the index dates. These medications have been identified as potential risk factors for kidney disease^7,16,36,37^. Drinking and smoking histories within 1 year before the index dates were also assessed based on ICD-10 codes and current procedural terminology (CPT) codes (*Supplementary Table 1*).

### Statistical Analysis

Age-adjusted demographic and clinical characteristics were summarized across the COVID-19 group, negative control group, and positive control group, with mean (SD) values for continuous variables, and number and percentage for categorical variables, which were presented after stratum matching. The incidence densities of AKI, CKD, and ESRD were then calculated per 1,000 person-years for the three groups. Additionally, cumulative incidence figures were presented to illustrate the trends of AKI, CKD, and ESRD incidence over the follow-up weeks across the three groups. Having confirmed no violations of the proportional hazards assumption (*Supplementary* Figure 1), we initially applied stratified Cox proportional hazards regression models, stratified by stratum-match ID, separately for overall kidney diseases and subgroups with CKD, AKI, and ESRD. Subsequently, multivariable stratified Cox proportional hazards regression models were performed, incorporating stratum-match ID, six comorbidities, seven drug use histories, drinking histories, and smoking histories. Given the reported complex combination effects between COVID-19 and each of diabetes^38^, CVD^39^, and chronic pulmonary disease^40^, interaction effects were assessed separately using the −2 × log likelihood ratio while controlling for the baseline covariates.

We conducted several sensitivity analyses to test the robustness of our results and to address the possibility of residual confounding: (1) All individuals from the 1536 strata were included in the analysis to address concerns regarding the exclusion of individuals without an exact 1:2 matching ratio. (2) Individuals without CVD, chronic pulmonary disease, diabetes, obesity, liver diseases, or depression before index dates were included to address the concern of residual confounding introduced from comorbidities. (3) Individuals who did not use ACEI/ARBs, statins, diuretics, CCB, NSAIDs, PPI, and beta-blockers were included, to address potential residual confounding from medications. (4) Propensity score strata were constructed using a logistic regression model with the covariates from the full model to balance baseline data between the COVID-19 group and the negative control group. Stratified Cox proportional hazards regression models were then conducted using the propensity score strata ID. (5) Individuals with index dates from January to June 2020 were included to address concerns regarding the unbalanced distribution of individuals in the influenza group.

We conducted an exploratory analysis to assess the risk of COVID-19 and influenza on glomerular diseases. Individuals who previously had glomerular diseases before the index dates were excluded from the analysis, and the same multivariable stratified Cox proportional hazards regression models were applied as the primary analysis.

Another exploratory analysis was performed to compare the effects of COVID-19 and influenza on AKI and CKD over 180, 360, and 540 days to assess effect trends with follow-up days. The same multivariable stratified Cox proportional hazards regression models used in the primary analysis were applied to these situations. ESRD and glomerular diseases were not included due to the limited number of incident cases during each follow-up period.

The final exploratory analysis was conducted to directly compare the hazards between COVID-19 and influenza, using influenza as the reference.

Data were analyzed in SAS software version 9.4 (SAS Institute, Cary, North Carolina) and R software version 3.6.2 (R Foundation for Statistical Computing, Vienna, Austria) using a two-tailed α level of 0.05. This study followed the Strengthening the Reporting of Observational Studies in Epidemiology (STROBE) reporting guideline for cohort studies^41^.

## Results

Our study included a total of 3,016,794 individuals, with a median follow-up time of 324 days (interquartile range: 186 to 429 days), divided into three groups: the COVID-19 group (n = 939,241; mean [SD] age 41.3 [13.1] years; 47.3% male), the negative control group (n = 1,878,482; mean [SD] age 41.3 [13.1] years; 47.3% male), and the influenza group (n = 199,071; mean [SD] age 39.7 [12.8] years; 43.1% male). After stratum matching, the COVID-19 and negative control groups showed identical distributions of age, gender, region, and index year. The influenza group showed a similar distribution to the negative control group regarding demographic information, except for the index year, with 92.8% of cases occurring between January and June 2020. To address potential biases introduced by the inconsistent distribution of index years, we conducted sensitivity analyses. Overall, the COVID-19 and influenza groups demonstrated a higher prevalence of comorbidities and medication use than the negative control group (*Table 1*).

**Table 1.** Age-adjusted demographic and clinical characteristics stratified by COVID-19 and Influenza status after stratum matching.

|  | COVID-19<br>(N= 939,241) | Negative Control<br>(N=1,878,482) | Influenza<br>(Positive Control)<br>(N= 199,071) |
| --- | --- | --- | --- |
| Age*, mean (SD) | 41.3 (13.1) | 41.3 (13.1) | 39.7 (12.8) |
| Gender |  |  |  |
| Male | 443,991 (47.3%) | 887,983 (47.3%) | 85,874 (43.1%) |
| Region |  |  |  |
| Northeast | 150,159 (16.0%) | 300,319 (16.0%) | 38,352 (19.3%) |
| North Central | 185,136 (19.7%) | 370,272 (19.7%) | 35,684 (17.9%) |
| South | 485,969 (51.7%) | 971,938 (51.7%) | 106,190 (53.3%) |
| West | 117,977 (12.6%) | 235,954 (12.6%) | 18,845 (9.5%) |
| Index Year |  |  |  |
| Jan. - June 2020 | 88,924 (9.5%) | 177,847 (9.5%) | 184,764 (92.8%) |
| July - Dec. 2020 | 367,195 (39.1%) | 734,390 (39.1%) | 5,977 (3.0%) |
| Jan. - June 2021 | 303,434 (32.3%) | 606,867 (32.3%) | 6,134 (3.1%) |
| July - Dec. 2021 | 179,689 (19.1%) | 359,377 (19.1%) | 2,196 (1.1%) |
| Comorbidities |  |  |  |
| Cardiovascular Disease, Yes (%) | 247,914 (26.4%) | 395,381 (21.0%) | 49,401 (24.8%) |
| Chronic Pulmonary Disease, Yes (%) | 75,568 (8.0%) | 105,710 (5.6%) | 20,351 (10.2%) |
| Diabetes, Yes (%) | 90,520 (9.6%) | 125,514 (6.7%) | 16,067 (8.1%) |
| Obesity, Yes (%) | 134,615 (14.3%) | 189,611 (10.1%) | 26,266 (13.2%) |
| Liver disease, Yes (%) | 27,509 (2.9%) | 38,772 (2.1%) | 5,498 (2.8%) |
| Depression, Yes (%) | 112,614 (12.0%) | 193,466 (10.3%) | 26,197 (13.2%) |
| Smoking, Yes (%) | 48,003 (5.1%) | 86,104 (4.6%) | 13,531 (6.8%) |
| Drinking, Yes (%) | 9,418 (1.0%) | 17,083 (0.9%) | 2,119 (1.1%) |
| Medications |  |  |  |
| ACEI/ARBs, Yes (%) | 143,810 (15.3%) | 251,743 (13.4%) | 27,256 (13.7%) |
| Beta-blockers, Yes (%) | 39,876 (4.2%) | 70,416 (3.7%) | 8,059 (4.0%) |
| CCB, Yes (%) | 62,494 (6.7%) | 108,784 (5.8%) | 11,815 (5.9%) |
| Diuretics, Yes (%) | 102,904 (11.0%) | 173,600 (9.2%) | 19,974 (10.0%) |
| NSAID, Yes (%) | 153,588 (16.4%) | 238,015 (12.7%) | 34,283 (17.2%) |
| PPIs, Yes (%) | 98,615 (10.5%) | 142,827 (7.6%) | 20,572 (10.3%) |
| Statins, Yes (%) | 123,213 (13.1%) | 227,654 (12.1%) | 23,673 (11.9%) |

\* Value is not age-adjusted; Binary and Categorical variables are standardized to the age distribution of the study population;
Values of Categorical variables may not sum to 100% due to rounding.
ACEI/ARBs: angiotensin-converting enzyme inhibitors or angiotensin II receptor blockers
CCB: calcium channel blockers
NSAID: nonsteroidal anti-inflammatory drugs
PPIs: proton pump inhibitors

COVID-19 group showed the highest incidence density in AKI (8.93 cases per 1000 PY, 95% CI (8.73, 9.14)), CKD (6.36 cases per 1000 PY, 95% CI (6.19, 6.54)), and ESRD (0.64 cases per 1000 PY, 95% CI (0.58, 0.69)), compared to negative control and influenza groups (*Figure 2*). In the cumulative incidence plots, COVID-19 group consistently exhibited the significantly highest cumulative incidence for AKI, CKD, and ESRD throughout the entire follow-up period. (*Figure 3*).

**Figure 2:**
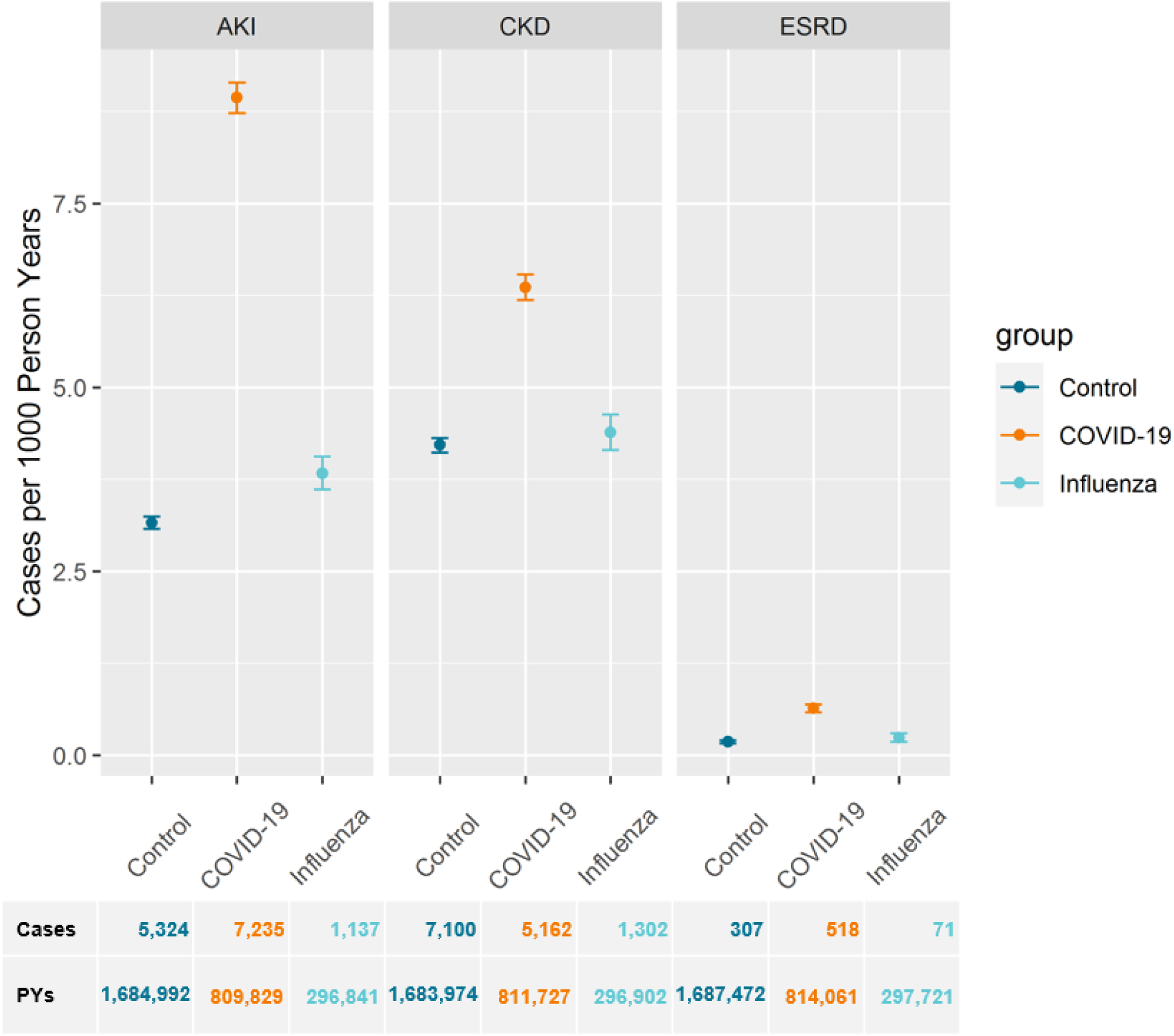
Incidence density of (1) Acute Kidney Injury (2) Chronic Kidney Disease (3) End Stage Renal Diseases across COVID-19, Influenza, and Negative Control groups (per 1000 person-years)

**Figure 3:**
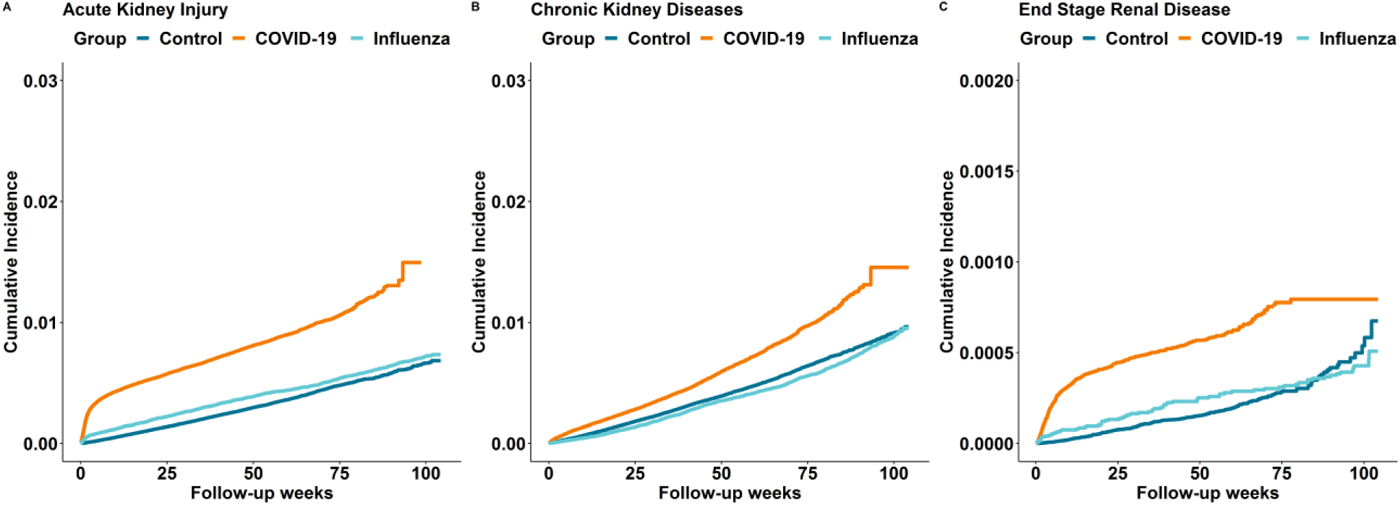
Cumulative Incidence of (1) Acute Kidney Injury (2) Chronic Kidney Disease (3) End Stage Renal Diseases across COVID-19, Influenza, and Negative Control groups

Stratified Cox proportional hazards regression models, adjusted for (1) match ID only and (2) match ID along with other baseline covariates, presented consistent results for the adjusted hazard ratios (aHR) for COVID-19 and influenza, using the negative control as the reference. Moreover, COVID-19 exhibited higher hazard ratios than influenza for both overall kidney diseases and subgroups of CKD, AKI, and ESRD. Specifically, after adjusting for baseline covariates, COVID-19 significantly increased risks of any kidney diseases (aHR, 1.93; 95% CI, 1.87 – 2.00), CKD (aHR, 1.38; 95% CI, 1.32 – 1.45), AKI (aHR, 2.74; 95% CI, 2.61 – 2.87), and ESRD (aHR, 3.22; 95% CI, 2.67 – 3.88). In contrast, influenza significantly but slightly increased the risks of any kidney disease when compared to negative control (aHR, 1.10; 95% CI, 1.01–1.19) and AKI (aHR, 1.24; 95% CI, 1.11–1.38), but had non-significant effects on incident CKD (aHR, 1.03; 95% CI, 0.92–1.14) and ESRD (aHR, 0.84; 95% CI, 0.55–1.29) (*Table 2*).

**Table 2.** Stratified Cox proportional hazards models HR (95% CI) for the association between COVID, Influenza, and kidney diseases.

| Models | Diseases | Negative Control | COVID-19 | Influenza |
| --- | --- | --- | --- | --- |
| Model 1 | Any Kidney Diseases | Reference | 2.08 (2.02, 2.13) | 1.17 (1.10, 1.24) |
|  | Chronic Kidney Disease | Reference | 1.54 (1.48, 1.60) | 1.09 (1.01, 1.17) |
|  | Acute Kidney Injury | Reference | 2.87 (2.77, 2.97) | 1.30 (1.20, 1.40) |
|  | End Stage Renal Disease | Reference | 3.66 (3.18, 4.23) | 0.99 (0.73, 1.33) |
| Model 2 | Any Kidney Diseases | Reference | 1.93 (1.87, 2.00) | 1.10 (1.01, 1.19) |
|  | Chronic Kidney Disease | Reference | 1.38 (1.32, 1.45) | 1.03 (0.92, 1.14) |
|  | Acute Kidney Injury | Reference | 2.74 (2.61, 2.87) | 1.24 (1.11, 1.38) |
|  | End Stage Renal Disease | Reference | 3.22 (2.67, 3.88) | 0.84 (0.55, 1.29) |

Model 1: Stratified with match ID
Model 2: Stratified with match ID, 6 comorbidities (cardiovascular disease, chronic pulmonary disease, diabetes, obesity, liver diseases, depression), 7 drug use histories (angiotensin-converting enzyme inhibitors or angiotensin II receptor blockers (ACEI/ARBs), statins, diuretics, calcium channel blockers (CCB), nonsteroidal anti-inflammatory drugs (NSAIDs), proton pump inhibitors (PPIs), and beta-blockers), drink histories, and smoking histories.

Interaction effects of COVID-19 with diabetes, CVD, and chronic pulmonary diseases were tested and none were statistically significant (all p > 0.05) (*Supplementary Table 2*). The results of sensitivity analyses in five different scenarios are consistent with the conclusions drawn from our primary analysis (*Table 3*).

**Table 3.**
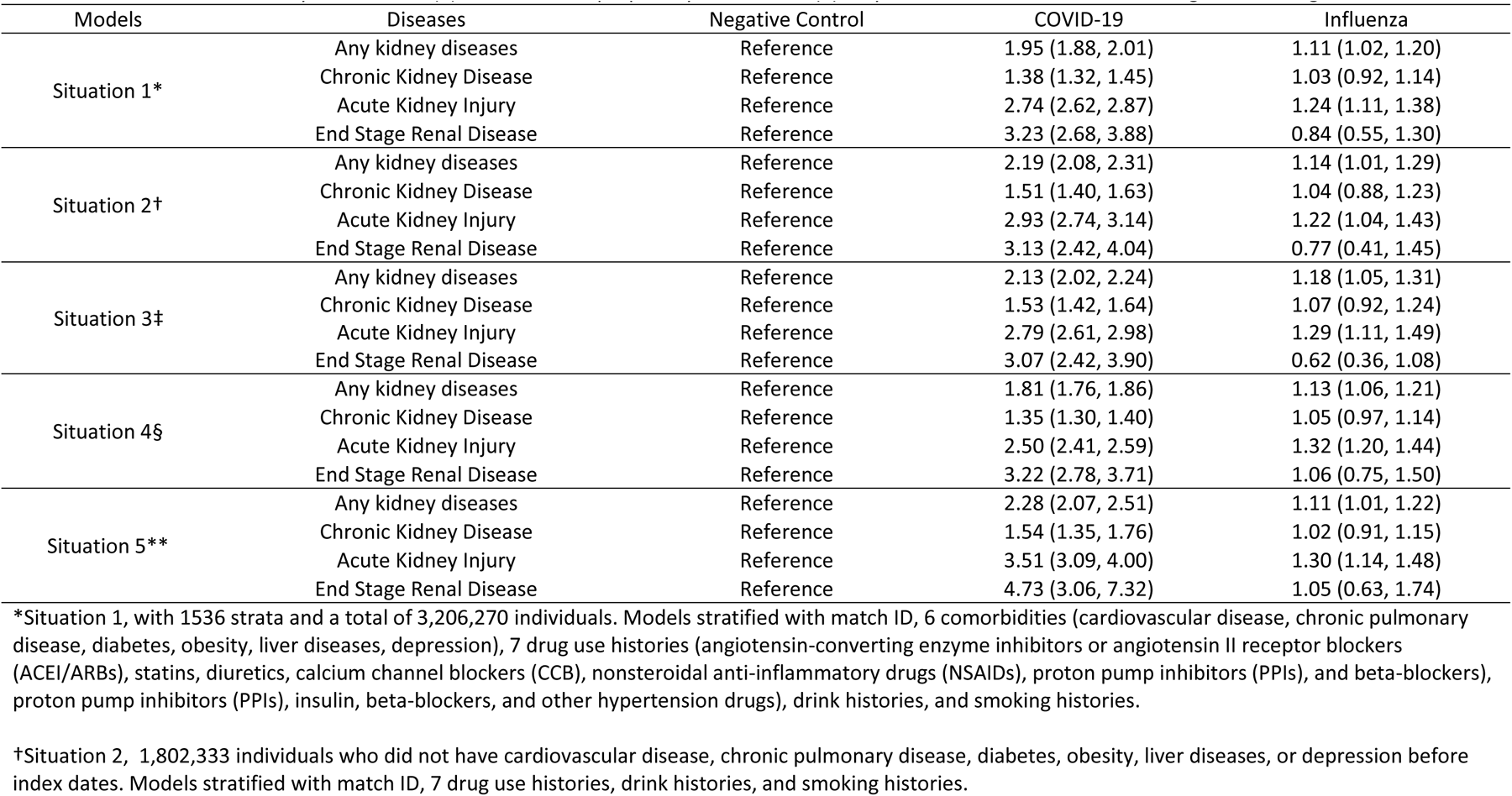
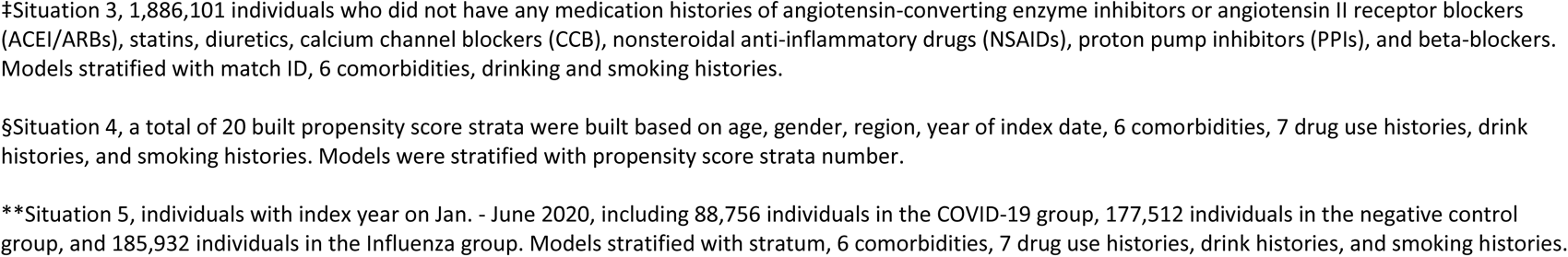
Sensitivity analysis for the association between COVID, Influenza, and kidney diseases: (1) all strata (2) exclude individuals who had any comorbidities (3) exclude individuals who had any medications (4) stratified with propensity score strata (5) only include individuals who were diagnosed during Jan. to June 2020.

| Models | Diseases | Negative Control | COVID-19 | Influenza |
| --- | --- | --- | --- | --- |
| Situation 1* | Any kidney diseases | Reference | 1.95 (1.88, 2.01) | 1.11 (1.02, 1.20) |
|  | Chronic Kidney Disease | Reference | 1.38 (1.32, 1.45) | 1.03 (0.92, 1.14) |
|  | Acute Kidney Injury | Reference | 2.74 (2.62, 2.87) | 1.24 (1.11, 1.38) |
|  | End Stage Renal Disease | Reference | 3.23 (2.68, 3.88) | 0.84 (0.55, 1.30) |
| Situation 2† | Any kidney diseases | Reference | 2.19 (2.08, 2.31) | 1.14 (1.01, 1.29) |
|  | Chronic Kidney Disease | Reference | 1.51 (1.40, 1.63) | 1.04 (0.88, 1.23) |
|  | Acute Kidney Injury | Reference | 2.93 (2.74, 3.14) | 1.22 (1.04, 1.43) |
|  | End Stage Renal Disease | Reference | 3.13 (2.42, 4.04) | 0.77 (0.41, 1.45) |
| Situation 3‡ | Any kidney diseases | Reference | 2.13 (2.02, 2.24) | 1.18 (1.05, 1.31) |
|  | Chronic Kidney Disease | Reference | 1.53 (1.42, 1.64) | 1.07 (0.92, 1.24) |
|  | Acute Kidney Injury | Reference | 2.79 (2.61, 2.98) | 1.29 (1.11, 1.49) |
|  | End Stage Renal Disease | Reference | 3.07 (2.42, 3.90) | 0.62 (0.36, 1.08) |
| Situation 4§ | Any kidney diseases | Reference | 1.81 (1.76, 1.86) | 1.13 (1.06, 1.21) |
|  | Chronic Kidney Disease | Reference | 1.35 (1.30, 1.40) | 1.05 (0.97, 1.14) |
|  | Acute Kidney Injury | Reference | 2.50 (2.41, 2.59) | 1.32 (1.20, 1.44) |
|  | End Stage Renal Disease | Reference | 3.22 (2.78, 3.71) | 1.06 (0.75, 1.50) |
| Situation 5** | Any kidney diseases | Reference | 2.28 (2.07, 2.51) | 1.11 (1.01, 1.22) |
|  | Chronic Kidney Disease | Reference | 1.54 (1.35, 1.76) | 1.02 (0.91, 1.15) |
|  | Acute Kidney Injury | Reference | 3.51 (3.09, 4.00) | 1.30 (1.14, 1.48) |
|  | End Stage Renal Disease | Reference | 4.73 (3.06, 7.32) | 1.05 (0.63, 1.74) |

\*Situation 1, with 1536 strata and a total of 3,206,270 individuals. Models stratified with match ID, 6 comorbidities (cardiovascular disease, chronic pulmonary disease, diabetes, obesity, liver diseases, depression), 7 drug use histories (angiotensin-converting enzyme inhibitors or angiotensin II receptor blockers (ACEI/ARBs), statins, diuretics, calcium channel blockers (CCB), nonsteroidal anti-inflammatory drugs (NSAIDs), proton pump inhibitors (PPIs), and beta-blockers), proton pump inhibitors (PPIs), insulin, beta-blockers, and other hypertension drugs), drink histories, and smoking histories.
†Situation 2, 1,802,333 individuals who did not have cardiovascular disease, chronic pulmonary disease, diabetes, obesity, liver diseases, or depression before index dates. Models stratified with match ID, 7 drug use histories, drink histories, and smoking histories.
‡Situation 3, 1,886,101 individuals who did not have any medication histories of angiotensin-converting enzyme inhibitors or angiotensin II receptor blockers (ACEI/ARBs), statins, diuretics, calcium channel blockers (CCB), nonsteroidal anti-inflammatory drugs (NSAIDs), proton pump inhibitors (PPIs), and beta-blockers. Models stratified with match ID, 6 comorbidities, drinking and smoking histories.
§Situation 4, a total of 20 built propensity score strata were built based on age, gender, region, year of index date, 6 comorbidities, 7 drug use histories, drink histories, and smoking histories. Models were stratified with propensity score strata number.
\*\*Situation 5, individuals with index year on Jan. - June 2020, including 88,756 individuals in the COVID-19 group, 177,512 individuals in the negative control group, and 185,932 individuals in the Influenza group. Models stratified with stratum, 6 comorbidities, 7 drug use histories, drink histories, and smoking histories.

For the exploratory analysis of glomerular disease, compared to the negative control group, after adjusting for baseline covariates, COVID-19 was associated with increased risk of glomerular disease (aHR 1.28, 95% CI 1.09–1.50), and influenza also elevated the risk (aHR 1.40, 95% CI 1.03–1.91).

In the exploratory analysis with follow-up periods, both COVID-19 and influenza showed significantly increased risks of AKI across 180, 360, and 540 days. However, COVID-19 (aHR, 4.35; 95% CI, 4.09– 4.63) had nearly three times the hazard ratio of influenza (aHR, 1.51; 95% CI, 1.29–1.78) for AKI within the first 180 days after infection. The impact on AKI declined for both COVID-19 (aHR, 2.75; 95% CI, 2.62–2.88) and influenza (aHR, 1.28; 95% CI, 1.14–1.44) by 540 days. Both COVID-19 and influenza showed relatively stable trends in their effects on CKD from 0–180 days to 0–540 days. However, COVID-19 showed significant impact on CKD across all three periods, while influenza remained non-significant (*Figure 4*).

**Figure 4:**
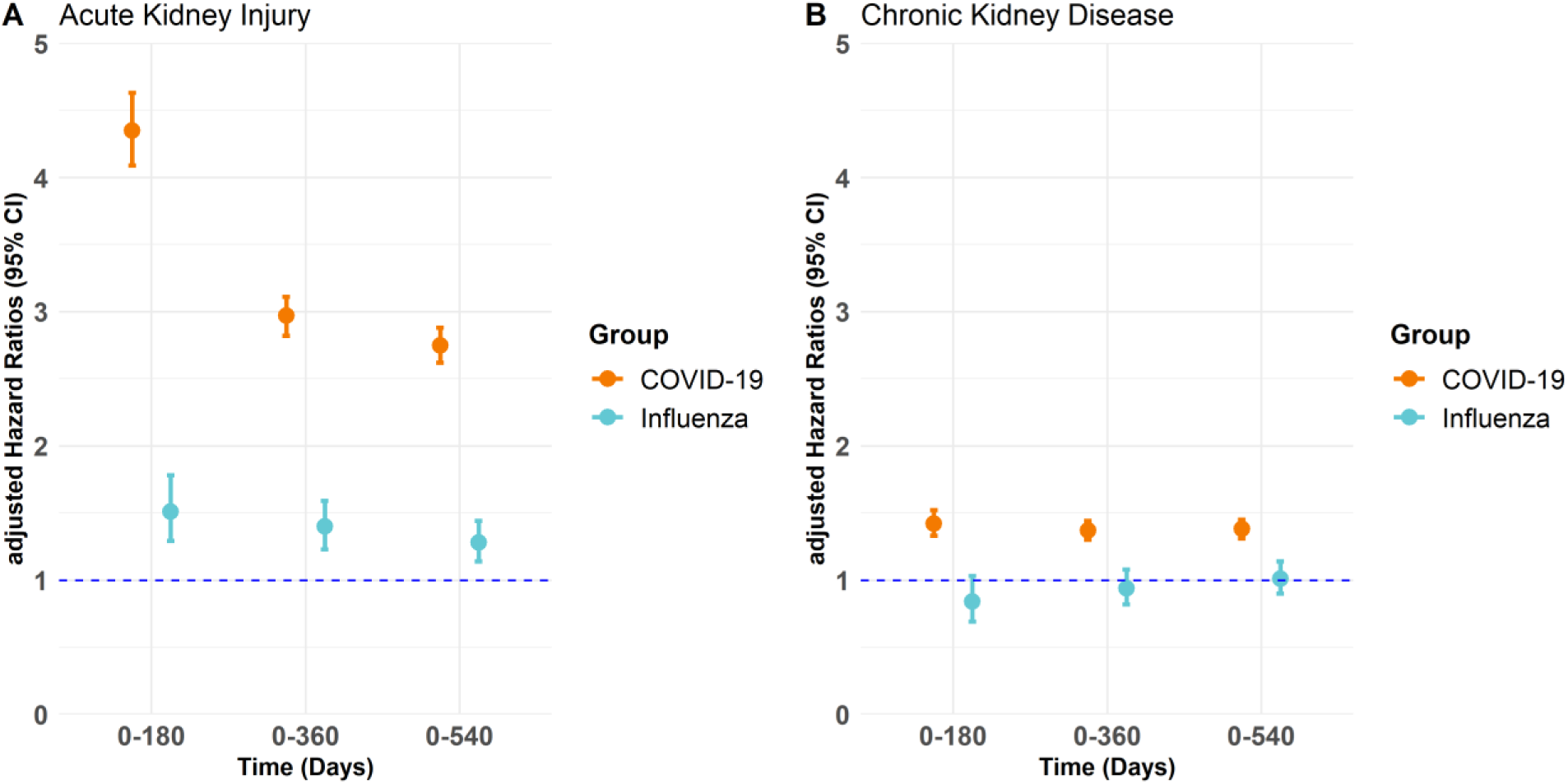
Exploratory Analysis of Chronic Kidney Disease and Acute Kidney Injury Risks Associated with COVID-19 and Influenza Compared to Negative Control Across Three Time Intervals: 0–180, 0–360, and 0–540 Days (Hazard Ratios, 95% Confidence Interval).

Exploratory analysis for the effects of COVID-19 with influenza as the positive control group showed significantly increased risks of AKI (aHR 2.28, 95% CI 2.00–2.60), CKD (aHR 1.39, 95% CI 1.21–1.60), and ESRD (aHR 4.67, 95% CI 2.85–7.66), but a non-significant impact on glomerular diseases (aHR 1.08, 95% CI 0.73–1.60) (*Figure 5*).

**Figure 5:**
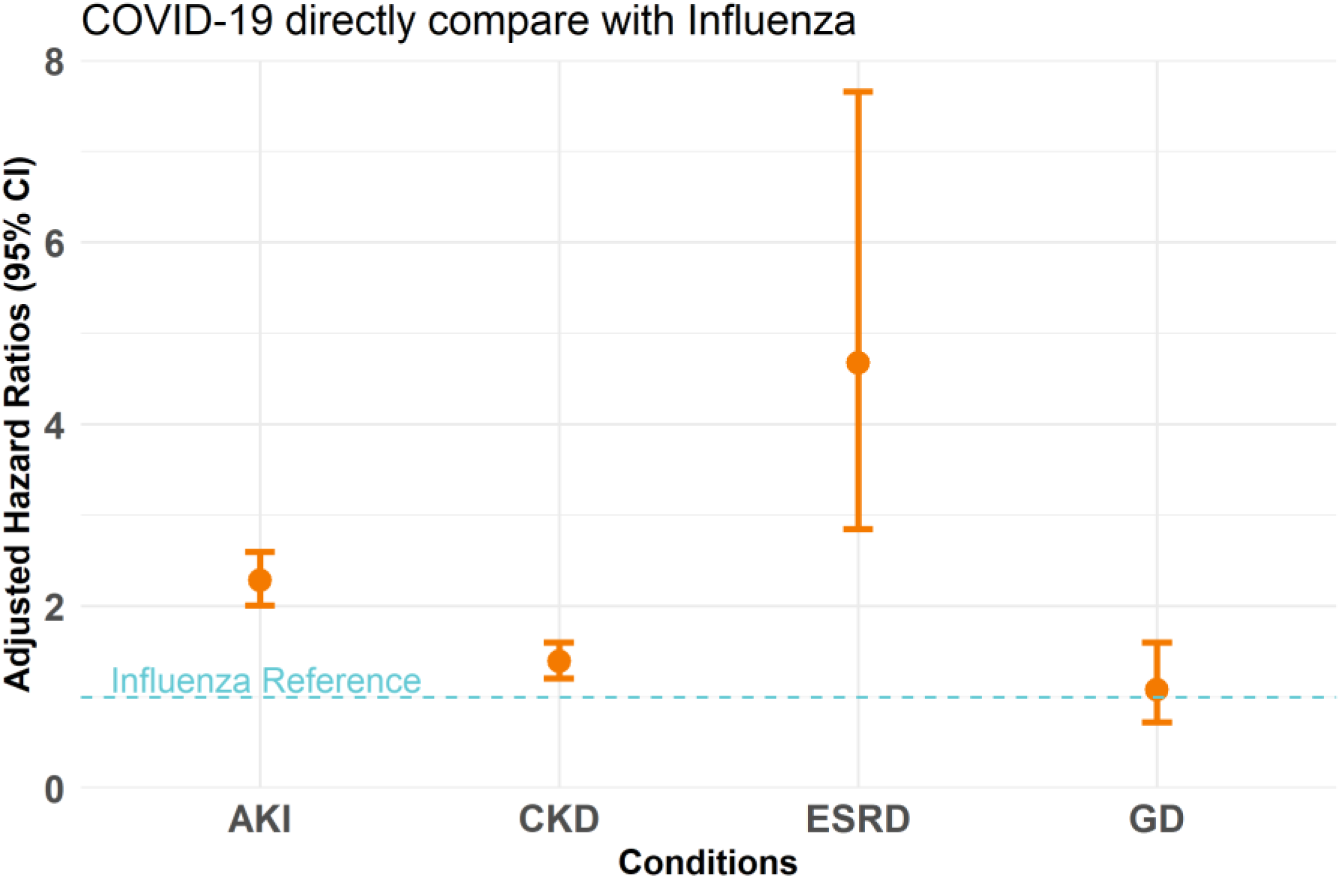
Exploratory Analysis of COVID-19 Impacts on Kidney Diseases Using Influenza as the Reference Group (Adjusted Hazard Ratios, 95% Confidence Interval)

## Discussion

In this large national retrospective cohort study of 3,016,794 individuals, we found that COVID-19 was associated with significantly higher risks of incident AKI, CKD, ESRD, and glomerular diseases, independent of baseline lifestyle factors and comorbidity conditions. In contrast, influenza was associated with a significant but mild effect on incident AKI and had no significant effects on CKD, ESRD, and glomerular diseases. Furthermore, COVID-19 had stronger effects on AKI within 180 days post-infection but demonstrated stable effects on CKD from 0-180 days to 0-540 days post-infection. Conversely, influenza had relatively stable mild effects on AKI after infection and showed no impact on CKD at any observed intervals. To our knowledge, our study is the first to comprehensively compare the associations of COVID-19 and influenza on AKI, CKD, and ESRD using large national real-world datasets. Moreover, this study is the first to explore the risk of COVID-19 on glomerular diseases based on real-world evidence.

Our findings are consistent with several previous studies. Bowe et al. conducted a retrospective cohort study using the Veterans Health Administration (VHA) healthcare system data to compare COVID-19 individuals with non-infected individuals from March 1, 2020, to March 15, 2021 (median follow-up days with 172 days). They found that the COVID-19 group had significantly increased risks of AKI (aHR 1.94, 95% CI 1.86 to 2.04) and ESRD (aHR 2.96, 95% CI 2.49 to 3.51)^7^. Similarly, our study reached the same conclusions but reported slightly higher aHR, likely due to the longer follow-up days in our study (median follow-up days with 324 days), and the different study population (91.2% of individuals in VHA are males). Xie et al. compared the risk of COVID-19 to influenza on 91 comorbidities in a retrospective cohort study using VHA data. They observed higher risks of AKI and CKD in the COVID-19 group compared to the influenza group^12^. Moreover, several prospective cohort studies with smaller sample sizes have observed increased risks of glomerular diseases (including proteinuria and hematuria) following COVID-19 infections, which are consistent with our exploratory results on glomerular diseases^42,43^.

Compared to influenza, COVID-19 has potential pathways leading to kidney diseases. First, Severe Acute Respiratory Syndrome Coronavirus-2 (SARS-CoV-2) enters kidney cells through specific biomechanisms. The virus binds to angiotensin-converting enzyme 2 (ACE2) via its spike (S) protein, which is then cleaved by the Type 2 transmembrane serine protease (TMPRSS2) or other proteases, forming a fusion pore^44^. Renal parenchymal cells, express high levels of ACE2, TMPRSS2, and other proteases, facilitating S protein cleavage and viral entry, making the kidneys susceptible to SARS-CoV-2 infection^9,45^. Moreover, several studies have found SARS-CoV-2 RNA and protein in the kidneys of COVID-19 individuals, which indicates the kidney is a target for SARS-CoV-2^46–48^. In contrast, although some studies have identified pathways for influenza A virus entry into host cells and its potential replication in human kidney cells^49^, further evidence is still needed^50^. Secondly, current observational studies have reported higher risks of comorbidities such as CVD and metabolic diseases in COVID-19 individuals compared to influenza^12,13^. These elevated comorbidity risks may also explain the increased incidence of kidney diseases in COVID-19 individuals compared to those with influenza^9,51^. Additionally, our study is the first to compare the effects of COVID-19 to influenza on glomerular diseases using large real-world data, providing evidence and valuable information for both mechanism and epidemiological research on glomerular diseases after COVID-19 infections.

Our study has multiple strengths. First, we employed comprehensive epidemiologic study designs to mitigate potential bias and confounding effects. We performed stratum matching to better balance the distribution of demographic characteristics and matched diagnosis dates to reduce bias from virus variants. Additionally, we included both positive and negative control groups to better evaluate the effects of COVID-19. In the negative control groups, we randomly assigned index dates for individuals, systematically excluding person-time before the random index dates, which may lead to a potential overestimation of the hazard ratio. By including influenza as a positive control group, with index dates corresponding to diagnosis dates, we avoided such overestimation when comparing COVID-19 directly with influenza.

This study also has some limitations. The MarketScan dataset is a claims dataset with insured individuals aged between 18 and 64 years. Our findings may not be generalized to elderly or pediatric populations, individuals without health insurance, and those covered by Medicare or Medicaid^52^. Additionally, MarketScan does not provide information on race or lab test results, preventing us from adjusting for potential unmeasured confounding by race and baseline creatinine levels. Additionally, although we included a large number of pre-defined covariates, such as diagnosed comorbidities and medications, and balanced these covariates between the COVID-19 groups and the control groups, we cannot completely rule out misclassification bias and residual confounding due to the nature of the observational study.

Lastly, we acknowledge the potential information bias in the diagnosis of kidney diseases, particularly mild CKD. The incidence of kidney diseases may be underestimated due to limited access to healthcare services, affecting both the COVID-19 and control groups, as restricted healthcare access can result in undiagnosed cases^53^.

## Conclusions

In our large national retrospective cohort study, COVID-19 was associated with a 1.28 times higher risk in AKI, 0.39 times higher risk in CKD, and 3.67 times higher risk in ESRD, compared to influenza.

COVID-19 has a stronger effect on AKI in the short term but may have stable long-term effects on CKD. Greater attention should be given to kidney disease in individuals with COVID-19 infections.

## Data Availability

Data are available from third-party partners Merative MarketScan Commercial Database

## Statements

### Ethics approval statement

The protocol of this study was reviewed and received a determination of non-human subjects’ research by the Penn State Institutional Review Board. The individual informed consent requirement was waived for this secondary analysis of de-identified data.

### Authors’ contributions

Designed research (project conception, development of overall research plan): YZ, DMB, NG, and VMC. Data extraction and study oversight: YZ, DMB. Analyzed data: YZ, and DMB. Performed statistical analysis: YZ. Wrote the first draft of the manuscript: YZ. Review and editing: YZ, NG, VMC, and DMB. All authors have read and approved the final manuscript. This study is part of the YZ’s doctoral dissertation research project with the Penn State College of Medicine, United States of America.

### Data sharing statement

Data are available from third-party partners Merative™ MarketScan^®^ Commercial Database

## Acknowledgments

None

