## Supplementary for "Does COVID-19 Increase the Risk of Subsequent Kidney Diseases More Than Influenza? A Retrospective Cohort Study Using Real-World Data In the United States"

**Supplementary Table 1:** Diagnosis codes of the International Classification of Diseases 10th editions, Clinical Modification (ICD-10-CM), used to describe baseline/preexisting clinical medical conditions

| **Variables** | **ICD-10 codes** |
|---|---|
| COVID-19 diagnosis | U07.1, B97.29, J12.82, B34.2, J12.81. |
| Influenza | J09, J10, J11 |
| Acute kidney injury | N17 |
| Chronic kidney disease | N18 |
| End-stage renal disease | Dialysis (Z99.2), Kidney transplant (Z94.0) |
| Glomerular diseases | N00-N08 |
| Congestive Heart Failure | I099, I110, I130, I132, I255, I420, I425, I426, I427, I428, I429, I43, I50, P290 |
| Cardiac arrhythmia | I441, I442, I443, I456, I459, I47, I48, I49, R000, R001, R008, T821, Z450, Z950 |
| Valvular disease | A520, I05, I06, I07, I08, I091, I098, I34, I35, I36, I37, I38, I39, Q230, Q231, Q232, Q233, Z952, Z953, Z954 |
| Pulmonary circulation disorders | I26, I27, I280, I288, I289 |
| Peripheral vascular disorders | I70, I71, I731, I738, I739, I771, I790, I792, K551, K558, K559, Z958, Z959 |
| Hypertension | I10, I11, I12, I13, I15 |
| Chronic Pulmonary Disease | I27.x, J40.x-J47.x, J60.xJ68.x, J70.x |
| Mild Liver Disease | B18.x, K70.x K71.x, K73.x, K74.x, K76.x, Z94.x |
| Diabetes | E10.x-E14.x |
| Obesity | E66 |
| Depression | F204, F313, F314, F315, F32, F33, F341, F412, F432 |
| Smoking | ICD-10: F17, Z716, Z720, T652, O9933, Z87891<br>CPT: 99406, 99407, G0375, G0376, G0436, G0437, G9016, G9276, G9458, G8402, G8403, G8453, G8454, S4990, S4991, S4995, S9075, S9453, 4000F, 4001F |

| | |
|---|---|
| Drinking | F10, K70, T51, G312, G621, G721, I426, K292, K852, K860, Y912, Y913 |

**Supplementary Table 2:** Hazard Ratios for Kidney Diseases Based on COVID-19 or Influenza Status, Stratified by Diabetes, Cardiovascular Disease, and Chronic Pulmonary Disease

| Variables | Negative Control | COVID-19 | Influenza | P-interaction |
|---|---|---|---|---|
| **Diabetes** | | | | 0.2192 |
| Yes | 1 (Ref.) | 1.82 (1.68, 1.98) | 1.16 (0.95, 1.43) | |
| No | 1 (Ref.) | 1.95 (1.88, 2.03) | 1.09 (1.00, 1.19) | |
| **Cardiovascular Disease** | | | | 0.5561 |
| Yes | 1 (Ref.) | 1.85 (1.68, 2.02) | 1.10 (0.89, 1.38) | |
| No | 1 (Ref.) | 1.94 (1.88, 2.02) | 1.10 (1.01, 1.20) | |
| **Chronic Pulmonary Disease** | | | | 0.0577 |
| Yes | 1 (Ref.) | 1.61 (1.35, 1.89) | 1.00 (0.72, 1.38) | |
| No | 1 (Ref.) | 1.95 (1.88, 2.02) | 1.11 (1.02, 1.20) | |

**Supplementary Figure 1:** log-log survival curves for checking the PH assumption for COVID-19 group and Negative Control groups were almost parallel and the proportional hazard assumption was satisfied

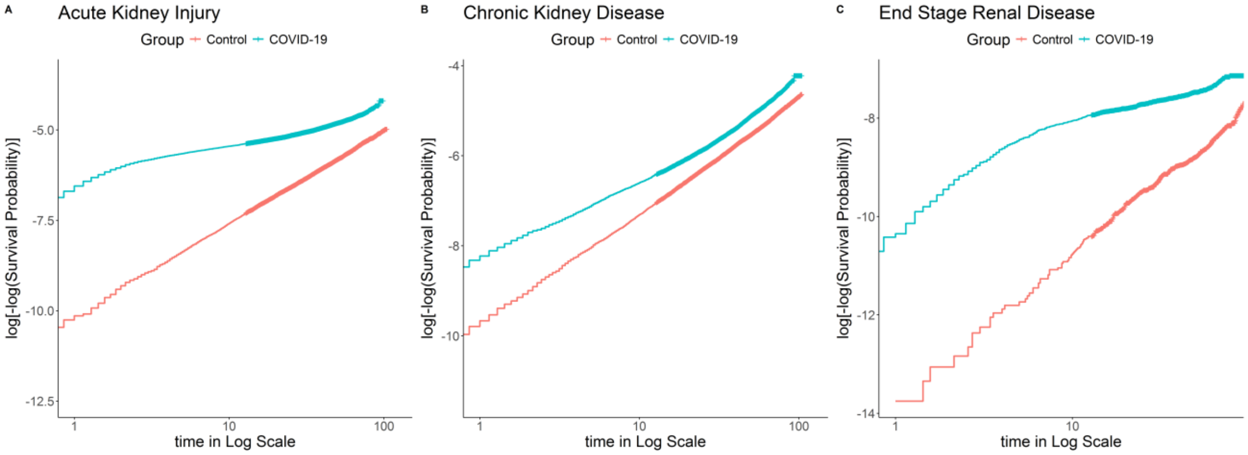
